# Fatal Iatrogenic Cerebral Amyloid-Related Encephalitis in a patient treated with lecanemab for Alzheimer’s disease: neuroimaging and neuropathology

**DOI:** 10.1101/2023.04.26.23289061

**Authors:** Elena Solopova, Wilber Romero-Fernandez, Hannah Harmsen, Lissa Ventura-Antunes, Emmeline Wang, Alena Shostak, Jose Maldonado, Manus Donahue, Daniel Schultz, Thomas M. Coyne, Andreas Charidimou, Matthew Schrag

## Abstract

We report the case of an elderly woman in good general health aside from early-stage Alzheimer’s disease who was enrolled in a randomized controlled trial of the novel therapeutic monoclonal antibody lecanemab where she was treated with the placebo, and subsequently in the open label extension study where she received lecanemab infusions every two weeks. After the third infusion, she suffered a seizure followed by aphasia and progressively worsening encephalopathy. Magnetic resonance imaging revealed multifocal cerebral edema and an increased burden of cerebral microhemorrhages compared to pre-trial imaging, consistent with Amyloid Related Imaging Abnormality (ARIA). She was treated with an antiepileptic regimen and high-dose intravenous corticosteroids but continued to worsen and expired after five days in the hospital. The family requested an autopsy and consented to evaluation of her brain for research. Post-mortem MRI confirmed extensive microhemorrhagic changes in the temporal, parietal and occipital lobes, some of which were visible on gross inspection of the brain. Autopsy confirmed APOE genotype of E4/E4 and the presence of typical neuropathological features of Alzheimer’s disease along with severe cerebral amyloid angiopathy with inflammatory features, including perivascular lymphocytic infiltrates, reactive macrophages and fibrinoid degeneration of vessel walls. There were deposits of β-amyloid in meningeal vessels and penetrating arterioles with numerous microaneurysms. Cerebral microhemorrhages were associated with arterioles harboring β-amyloid deposits and having degenerative morphologies. We conclude from these results that the patient likely died as a result of severe cerebral amyloid angiopathy with marked microvascular degeneration and meningoencephalitis. Further study of the mechanism of ARIA and the neuropathological changes associated with plaque clearance are needed.

## Introduction

Over the last 15 years, numerous clinical trial programs have tested infusions of monoclonal antibodies against various forms of β-amyloid in an effort to facilitate clearance of plaques, oligomers and/or soluble β-amyloid peptide from the brains of patients with Alzheimer’s disease. This approach emerged on the heels of a promising, although ultimately unsuccessful, effort to vaccinate patients with Alzheimer’s disease against β-amyloid, which ended in early 2002 after a subset of patients developed encephalitis (1). Neuropathologic characterization of several of those patients showed they had accelerated cerebral amyloid angiopathy leading to a breakdown of the blood brain barrier, inflammatory infiltration, and bleeding (2). The underlying pathophysiology of this side effect was felt to be due to rapid clearance of β-amyloid through the perivascular pathway overwhelming the efflux capability the microvascular network leading to injury. These findings in β-amyloid vaccination were similar to pathological features seen in the spontaneous disorder known as cerebral amyloid angiopathy related inflammation (CAARI) (which has sometimes been termed amyloid-β related angiitis or ABRA when the pathological features are particularly aggressive) (3). With passive antibody administration, a similar phenomenon has been observed and termed ARIA or β-amyloid related imaging abnormality. This side effect is common, occurring in up to 40% of patients in some trials, although most cases have been less severe than the encephalitis associated with β-amyloid vaccination (4). The neuropathological features of ARIA are not well-understood owing to few cases in the published literature, and consequently there is some debate whether the mechanism of ARIA is equivalent to the earlier encephalitis cases and to CAARI. Here we report the clinical, neuroimaging and neuropathological features of a lethal case of ARIA associated with infusions of the anti-β-amyloid monoclonal antibody lecanemab (originally known as BAN-2401). We conducted post-mortem neuroimaging and histological studies to evaluate the tissue changes associated with this syndrome.

### Brief case history

A woman between 75 and 80 years old in good general health, with no clinical history of hypertension or neurological disorder except for an approximately four-year history of mild but progressively worsening memory symptoms. Tau and β-amyloid PET scans supported the diagnosis of Alzheimer’s Disease, and she was enrolled in a randomized controlled trial of lecanemab where she was assigned to the placebo group. The patient reported no adverse effects nor perceptible benefits, and at the completion of the trial she was enrolled in the open-label extension which guaranteed treatment with the active drug. She was treated with three infusions of lecanemab, each dose (10 mg/kg) approximately two weeks apart. According to her study partner, approximately an hour after each dose she developed a headache causing her to spend one to two days in bed recovering each time. After the third dose, she began to experience progressively worsening memory impairment which she described as “brain fog.” On the day of hospital admission, she had a seizure which began with left head and gaze version and left sided tonic contraction which evolved into 30 seconds of generalized convulsion. She regained alertness after this event but was never communicative nor purposefully interactive again. Electroencephalography did not suggest ongoing seizures to explain her condition, although she had frontal intermittent rhythmic delta activity. Neuroimaging revealed cerebral edema in the bilateral temporal, parietal and occipital lobes with numerous microhemorrhages. She was treated with 1g daily intravenous solumedrol for three days for suspected amyloid-related imaging abnormality (ARIA) without improvement. She suffered an aspiration event leading to sepsis with multiorgan failure and, consistent with her wishes, aggressive resuscitation measures were not instituted. She expired 5 days after hospital admission. An extended case history is included in the supplementary material.

### Neuroimaging

A pre-treatment MRI was obtained as part of the enrollment procedure before the open-label extension component of the clinical trial and a post-treatment MRI was obtained as part of her clinical care after her seizure and neurological deterioration. The pre-treatment study showed moderate white matter disease without mass effect on FLAIR sequences (Figure 1a and Supplementary Figure 1). The post-treatment study shows expansion of the white matter signal abnormality with edema in both temporal lobes, parietal lobes, and occipital lobes (Figure 1b and Supplementary Figure 2). The pre-treatment study showed four small cerebral microhemorrhages on gradient echo-T2* (Figure 1c and supplementary Figure 3). The post-treatment study shows more than 30 microhemorrhages, all in a cortical or juxtacortical distribution and most within areas of edema (Figure 1d and Supplementary Figure 4). Selected images from the diffusion weighted and apparent diffusion coefficient (ADC) scans associated with her hospital admission are shown in Supplementary Figure 5; there was no major stroke. Before initiating treatment with lecanemab in the open label extension study, the patient underwent a β-amyloid PET scan (with Fluorbetaben) and tau PET scan. Representative images from these studies are shown in Supplementary Figure 6. Tracer retention suggested both amyloid and tau positivity at entry into the open label extension phase of the trial.

**Figure 1:**
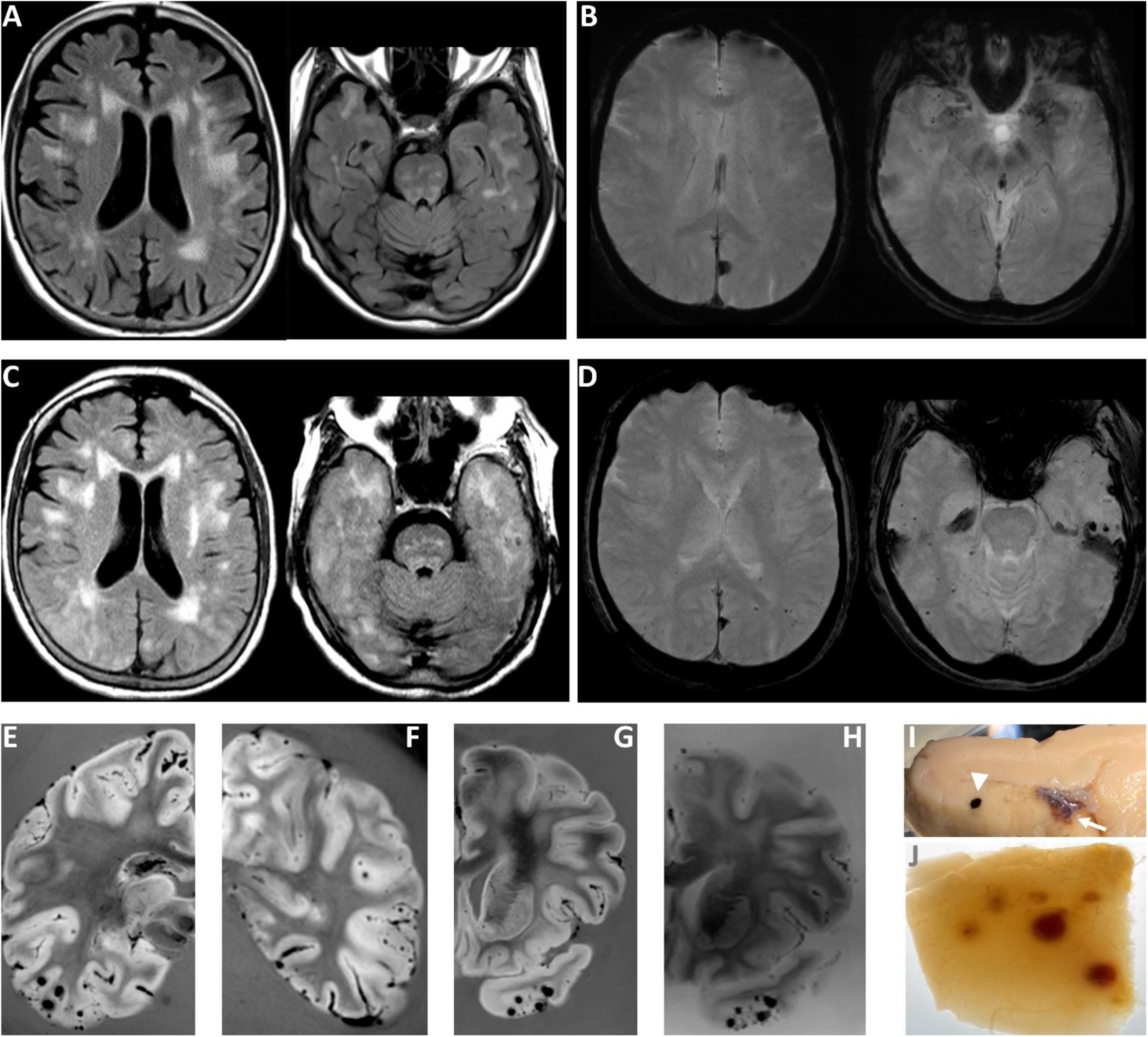
Neuroimaging evidence of cerebral edema and microhemorrhage A. Pre-trial FLAIR and B. susceptibility weighted (minimum intensity projection) images. Moderate pre-existing white matter disease was present along with four cortical microhemorrhages. C. Hospital-acquire FLAIR and D. susceptibility weighted (minimum intensity projection) images. Increased FLAIR hyperintensity and sulcal effacement/mass effect in the temporal, parietal and occipital lobes suggest the presence of edema. The number of microhemorrhages increased to over 30. E, F & G. Post-mortem susceptibility weighted imaging at 3T demonstrated extensive microhemorrhagic changes, most prominently in the temporal, parietal and occipital lobes. H. 7T imaging, paired with G shows that additional microhemorrhages are detected at high field strength. I. A few microhemorrhages (arrowhead) and superficial siderosis (arrow) were visible on gross sections. J. More extensive microhemorrhagic changes were readily observed when the tissue was rendered translucent with optical clearance approaches and backlighting.

The post-mortem brain was scanned separately at the clinical field strength of 3T and at the high field strength of 7T to further evaluate the extent of hemorrhagic changes in the brain (Figure 1e,f,g and h respectively). The hemorrhagic changes visualized on the post-mortem imaging are much more extensive than on the *in vivo* scan. To confirm the extent of microhemorrhagic changes, we subjected a 0.5 cm thick section of a gyrus from the temporal cortex to optical clearing and when it was translucent at roughly the midpoint during the clearance procedure, we photographed the block with backlighting to show the full extent of bleeding (Figure 1h). The density of microhemorrhages visualized pathologically and on post-mortem MRI is higher than was visualized on the clinical *in vivo* MRIs.

### Neuropathology

On gross inspection of the brain, there was evidence of edema in the temporal, parietal and occipital lobes based upon flattening of the cortical gyri against the leptomeninges. Petechial hemorrhages were visible on the natural and cut surfaces of the brain (Figure 2a-c). Sixteen blocks of tissue were prepared for histological evaluation (shown in Supplementary Figure 7). Immunohistochemistry for β-amyloid and phosphorylated tau demonstrated the presence of neuritic plaques and neurofibrillary tangles consistent with intermediate Alzheimer’s disease neuropathologic changes (Thal phase 5 of 5 for β-amyloid deposition, Braak and Braak stage IV of VI for neurofibrillary tangles, and CERAD frequent neuritic plaques: Alzheimer’s Disease Neuropathology Change A3, B2, C3). To further characterize the plaques, we stained sections with methoxy-XO4 (a sensitive blue-fluorescent probe for amyloid and aggregated tau), vacuolar ATPase subunit V0A1 or phospholipase D3 (two of the most sensitive markers of dystrophic neurites we have found to date)(5) and IBA1 (a marker of microglia). While most of the plaques looked entirely typical, 21% appeared to have been “cleared” – which is to say, a distinctive rosette of dystrophic neurites was present without the typical amyloid staining at its center. A further 24% of the plaques had minimal staining of amyloid deposits (Supplementary Figure 8). These features were associated in some cases with intense immunoreactivity for IBA1/microglia (Figure 2d). Numerous areas of microhemorrhage were visualized histologically along with arterioles with varying degrees of fibrinoid necrosis and perivascular inflammation. The bulk of the perivascular inflammation was composed of macrophages with occasional multinucleated giant cells; some T-cells were also present in the perivascular inflammatory milieu (Figure 2e-I and Supplementary Figure 9). The patient also had severe cerebral amyloid angiopathy. Culprit arterioles associated with microhemorrhages contained significant β-amyloid deposits (Figure 2j,k). Three-dimensional views of degenerating and ruptured arterioles were obtained from lightsheet microscopy of cleared tissue and are shown in the supplementary video. In many cases, blood extended from the site of arteriolar rupture within contiguous perivascular spaces, as was previously described (Figure 2k)(6). Vascular amyloid deposition and microaneurysms were also frequently observed in meningeal specimens (Figure 2l-n). APOE genotyping was not available from her clinical records, so this was obtained as part of her autopsy; she carried two copies of the E4 allele.

**Figure 2:**
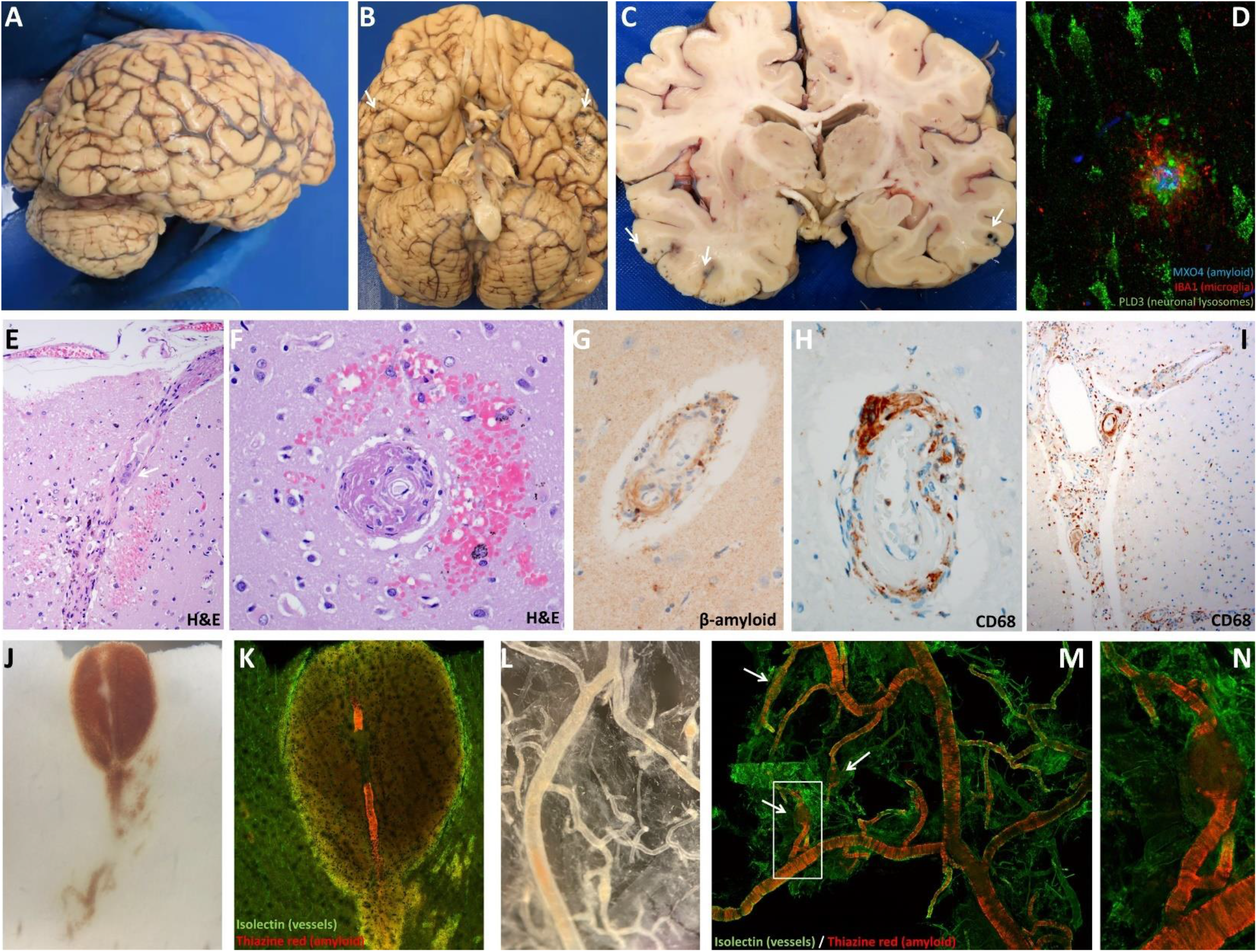
Inflammatory and microhemorrhagic pathology **A**. Lateral and **B**. inferior views of the brain. Black stippling in the temporal lobes consistent with petechial hemorrhage is best seen on the inferior view (arrows). **C**. A coronal section through the brain shows additional microhemorrhages. **D**. Methoxy-XO4 staining of amyloid (blue), phospholipase D3 (green, staining neuronal lysosomes and dystrophic neurites) and IBA1 (red, staining microglia) shows a neuritic plaque surrounded by dystrophic neurites and activated microglia. This case met criteria for moderate Alzheimer’s disease neuropathologic changes. Additional images of amyloid deposits in this case are shown in Supplementary Figure 8. **E**. Longitudinal and **F**. cross-sectional images through penetrating arterioles in the affected region shows intensive perivascular inflammatory infiltration and bleeding. A multinucleated giant cell is present at the arrow in F. **G**. Immunostaining for β-amyloid demonstrates inflamed vessels have β-amyloid deposition. **H**. Cross-sectional and **I**. longitudinal sections were immunostained for CD68, showing numerous macrophages on the inflamed vessels (see also Supplementary Figure 9). **J**. Serial sectioning through a microhemorrhage enabled visualization of the culprit ruptured vessel. **K**. The culprit vessel contains heavy β-amyloid deposits (stained red, vascular marker isolectin in green, blood in non-specific yellowish autofluorescence around the vessel). **L**. Meningeal whole mount specimen overlying temporal lobe shows several microaneurysms. **M**. Meningeal specimen from L stained with thiazine red for β-amyloid (red) and isolectin to label vessels (green) shows severe β-amyloid deposition, including in a number of microaneurysms (arrows, the aneurysm in the white box is shown enlarged in **N**).

## Discussion

This case shows the neuroimaging and neuropathological features of an acute cerebral vasculitis which occurred during lecanemab treatment, an experimental anti-βamyloid therapy for Alzheimer’s disease. The clinical, pathological and neuroimaging findings in this case are consistent with a severe form of a known side-effect of this class of drugs which has been termed amyloid related imaging abnormality, or ARIA (7). Neuropathologically, we found that her condition was associated with marked perivascular inflammation and arteriolar degeneration resembling fibrinoid necrosis, leading to microhemorrhagic changes both in the parenchyma and leptomeninges. The inflammatory features include extensive macrophages and/or activated microglia in the perivascular space, along with occasional multinucleated giant cells. These features were associated with severe cerebral amyloid angiopathy. This constellation of findings is similar to the sporadic condition cerebral amyloid angiopathy related inflammation. This case was refractory to treatment. In sporadic CAARI, most cases respond well to corticosteroid treatment, but a minority are refractory and may have poor outcomes.

This case highlights an important side effect of this class of drugs. Around 2010, the FDA recommended restricting patients with two or more microhemorrhages from participating in clinical trials of anti-amyloid immunotherapies due to the risk of developing a syndrome of vasogenic edema and/or microhemorrhages (7). An academic/industry collaborative working group was assembled by the Alzheimer’s Association to address this concern and advocate for less restriction in patient enrollment criteria (8, 9). This group introduced the term ARIA in 2011 to describe the syndrome, adding the modifier -E for edema or effusion and the modifier -H for the presence of hemorrhage (10). Although around 75% of patients who develop ARIA remain asymptomatic and most have a relatively benign course, severe cases like this one do occur. Because the neuropathological findings are quite similar to the cases of encephalitis after β-amyloid vaccination, we suggest the term iatrogenic cerebral amyloid related encephalitis (iCARE) be used when this syndrome is symptomatic (11, 12). The risk of developing iCARE correlates with the presence of CAA and associated risk factors, including APOE4 genotype (10, 13). It seems reasonable to avoid treating patients carrying two copies of the E4 genotype with this medication.

In this case, the patient met diagnostic criteria for CAA. Importantly, the *in vivo* MRI significantly underestimated the extent of microhemorrhages, confirmed by *post-mortem* imaging with optimized acquisition settings and on neuropathological inspection. Microhemorrhages on the clinical scan are certainly useful diagnostically but should be viewed as a marker of microvascular disease rather than an indicator of the extent of microvascular disease. The trial exclusion criteria permit enrollment of patients with up to 4 microhemorrhages. These criteria do not consider of the distribution of microhemorrhages and do not standardize the screening MRI protocols, so will likely lead to heterogeneity in the detection of CAA. Detailed MRI-based diagnostic criteria for CAA have been established which are likely to perform better than the coarse criteria employed as exclusion criteria in this and many other clinical trials (14, 15). The difficulty with reliable detection of CAA is likely to be magnified as this treatment moves to the community setting where less sensitive sequences like gradient echo T2* and lower field strength imaging are prevalent, which is likely to permit more patients with co-morbid CAA to meet inclusion criteria. Another important safety consideration is that while patient screening and treatment could be restricted to facilities with adequate and standardized diagnostic capabilities, the treatment of side effects is likely to occur in the community setting in many cases. At present, ARIA or iCARE has rarely been encountered outside of a clinical trial population, so community health care providers would likely benefit from education on the features and risks associated with this side effect.

This case demonstrates an important gap in our understanding of the molecular mechanisms of ARIA or iCARE. It would be tempting to interpret this inflammatory condition as an antibody-mediated autoimmune attack against β-amyloid in the cerebral microvasculature. However, ARIA and iCARE rates in the clinical trials with aducanumab were also quite high, despite reports that aducanumab preferentially binds parenchymal versus vascular β-amyloid (16). An alternative interpretation is that the induction of β-amyloid clearance mediated by these antibodies traffics much of the β-amyloid through the perivascular spaces, leading to periarteriolar inflammation that can worsen CAA. If this perivascular clearance overwhelms the capacity of the microvasculature, blood brain barrier dysfunction may ensue initiating an inflammatory cascade. This latter interpretation is supported by animal studies (17, 18). Defining and mitigating the molecular mechanism of this side effect may be key to improving the safety of anti-β-amyloid therapies.

Additionally, it is not clear how the neuropathological features of Alzheimer’s disease change with β-amyloid lowering therapies. The available pre-clinical systems used to test β-amyloid-lowering therapies do not recapitulate all of the features of Alzheimer’s disease, so careful neuropathological study of patients who have been responsive to β-amyloid lowering therapies is critically needed. It will be important to determine whether dystrophic neurites resolve or are persistent, whether the various tau pathologies appear to be halted, whether granulovacuolar degenerating bodies remain and whether brain atrophy is reduced. Neuroimaging results would suggest brain atrophy is not halted (at least acutely) (19). These features will have considerable bearing on whether minor alterations in the cognitive trajectory of treated patients are likely to represent disease modification or purely symptomatic effects.

In conclusion, this case should prompt a careful evaluation of the safety of lecanemab treatment and a refinement of the approach to pre-screening potential recipients of lecanemab for cerebral amyloid angiopathy and its risk factors. We advocate that patients should be screened for cerebral amyloid angiopathy using the Boston criteria and should undergo APOE genotyping prior to treatment. The case also highlights an urgent need for neuropathological studies to address the features of ARIA and iCARE.

## Data Availability

All relevant data are included in this article and the associated supplemental material.

## Acknowledgements

We are grateful to the family of this patient who contributed her tissue and medical records for scientific study. The patient’s surrogates provided informed consent to participate in this case report. This study was IRB approved as part of our ongoing Observational Study of CAA and Related Disorders (OSCAAR), approval number 180287. This case study was made possible by philanthropic support from the family and friends of Louis Stephen Zuzga Moran and the family and friends of Douglas B. Janney, Jr.

MJD receives research related support from Philips North America; is a paid consultant for Pfizer Inc, Alterity, Global Blood Therapeutics, Graphite Bio, and LymphaTouch; is a paid advisory board member for Novartis and Bluebird Bio; receives research funding from Pfizer Inc; and is the President/CEO of Biosight, LLC which operates as a clinical research organization. AC reports receiving funding from the Bodossaki Foundation and the Frechette Family Foundation and consulting fees from Imperative Care. MSS reports receiving funding from the American Federation of Aging Research, the National Institutes of Health and consulting fees from Labaton-Sucharow LLP and Raymond James. The other authors declare no competing interests.

## Supplementary information

### 1. Expanded case history

#### Initial presentation

The patient was between 75 and 80 years old and in good general health prior to the admission in question; she had early-stage Alzheimer’s disease, no history of hypertension and only took medication for mild depression and gastroesophageal reflux. The patient’s study partner reported that she had been complaining of headaches and “brain fog” for the last several weeks; the headaches were most severe immediately after infusions of an experimental drug in a clinical trial and lasted one to two days during which she mostly stayed in bed. Her study partner additionally reported that in the days prior to the precipitating event she was more easily disoriented than typical. On the day in question, she was brought by EMS to the hospital for what her study partner described as a seizure. While eating at a restaurant, she looked and turned to the left, became stiff and her speech became slurred. She then had 30 seconds of generalized convulsion after which she slumped out of her chair due to flaccid left hemiplegia. She was intubated in the field for hypoxia (SpO2 ∼80%). Upon arrival in the ER her blood pressure was 126/99, rapid glucose screen was normal. An emergent CT scan of the head was interpreted as: “extensive low-density within the periventricular region bilaterally; no intracranial hemorrhage, mass or acute territorial infarcts.” The treating team initially felt her presentation was most worrisome for a stroke. The ER team and consulting neurologist considered administering a thrombolytic agent but decided to forego this treatment based on the unclear history and suboptimal neurological exam due to sedation from the recent intubation. The family informed the treating team that the patient “was getting some experimental medication for dementia at a research center -- she got 3 doses in the last six weeks.”

#### Medical history

She had mild dementia, but was independent, living with a roommate who was also her study partner. She participated in a randomized trial of an anti-β-amyloid immunotherapy called lecanemab for approximately 18 months (it has not been disclosed if she was receiving the placebo or active drug). She felt no obvious side effects nor obvious benefits during the randomized portion of the study. She recently enrolled in the open-label extension of the trial where she was guaranteed to receive the active drug and had received three infusions (10mg/kg) at two-week intervals. Preconditions of the study included documentation of pathological levels of β-amyloid by PET imaging; her imaging prior to enrollment in the open label extension continued to show amyloid positivity. Her other medical problems were gastroesophageal reflux, right knee replacement and a small meningioma which had been stable in size for decades. She notably had no prior history of seizures, no tobacco, alcohol, or sedative use.

#### Examination

Her initial examination upon arrival to the hospital was limited due to recent intubation, but once admitted to the ICU, she was unable to follow commands, and was agitated, but regained motor function on the left as she was noted to withdraw from noxious stimulation symmetrically in all extremities. She had no obvious cranial nerve defect.

#### Diagnostic evaluations

Initial labs were notable for leukocytosis (22,100/mm^3^ with a 92% neutrophilic predominance) which returned to nearly normal within 24 hours. Platelet count was normal (205,000/mm^3^), and she was not anemic (Hgb 12.8 g/dL). A chemistry panel showed normal renal function and normal liver function tests, but she developed elevated transaminases on hospital day 2 (AST 109 IU/L, ALT 78 IU/L, alkaline phosphatase 101 U/L, and total bilirubin 0.8 mg/dL). All sputum, blood and urine cultures were negative during the hospitalization.

Her EKG showed she was in atrial fibrillation. Echocardiogram was normal (EF 55%, normal valves, wall motion and chamber dimensions).

Her apolipoprotein genotype was evaluated in the course of the clinical trial, but the result has not yet been released.

EEG interpreted as follows: No posterior dominant rhythm; background consisted of 4-5 hz activity that appeared symmetric. There were frontal sharp waves that appeared rhythmic consistent with a FIRDA pattern. Photic stimulation evoked a driving response at certain flash frequencies.

The key sequences from the MRI are available in the attached imaging summary. Post-contrast images were also obtained (not shown) and were uninformative.

A baseline MRI was obtained before enrollment in the open label extension of the clinical trial (a little more than six weeks prior to her acute event) and is included in this material for additional context.

#### Hospital course

She was started on heparin infusion after atrial fibrillation was identified. Anticoagulation was monitored with factor Xa levels – the highest was 2.5x the upper limit of the reference range. The patient’s clinical trial physician raised a concern for the possibility of amyloid-related imaging abnormality (ARIA) as a side effect from the experimental drug. She was started on Keppra 500 mg BID and 1 g daily of solumedrol which was continued for four doses over three days. To manage agitation, she was sedated using variably fentanyl, propofol, midazolam and dexmedetomidine infusions. On day 2, she required diltiazem to control tachycardia and she developed an elevated highly sensitive troponin level (504 pg/ml). On the third day, the patient was extubated with instructions not to reintubate, in line with requests in her living will. She remained “agitated and restless” according to nursing staff, mute aside from occasional unintelligible mutterings, she was unable to follow commands, but spontaneously moved all extremities. Repeat CT head showed neither worsening nor improvement in the white matter hypodensities and no new pathology. At the end of hospital day 3, her breathing deteriorated and she became more hypoxic. A loop diuretic was administered, and she was placed on high-flow oxygen with bilevel positive airway pressure. Approximately 36 hours later, she had an aspiration event, developed respiratory distress and hypotension, and subsequently developed renal injury (Cr 2.1 mg/dL, BUN 48 mg/dL), leukocytosis (20,000/mm^3^), and worsened transaminitis (AST 2398 IU/L, ALT 1854 IU/L, alkaline phosphatase 187 U/L, tBili 2.4 mg/dL). Consistent with her wishes to forego invasive life support measures, the family and treating team instituted comfort-oriented care measures and the patient expired in about one day.

#### Autopsy

The brain was removed and immersion fixed in 10% formaldehyde prior to coronal sectioning into roughly 1 cm coronal slabs. For histological analysis, 16 blocks of tissue were paraffin embedded. The locations of the tissue blocks is shown in supplementary figure 7 and include the right middle frontal gyrus, right anterior superior and middle temporal gyrus, the right mid-to-posterior middle and inferior temporal gyri, the left striate region, the right anterior cingulate gyrus with corpus callosum, the right striatum, right basal ganglia at anterior commissure, right thalamus, left mid hippocampus, left amygdala, caudal midbrain/rostral pons, pons, medulla, left cerebellar cortex with dentate nucleus, right parietal cortex (with hemorrhage), and right inferior temporo-parietal cortex with hemorrhages. Microscopic neuropathological analysis was conducted independently by Dr. Coyne and Dr. Harmsen. The results were discussed by consensus conference.

On gross evaluation, the left cerebral hemisphere was mildly full in comparison to the right, and the cortical gyri were widened and flattened against the leptomeninges, consistent with moderate cerebral cortical edema. The large blood vessels arose normally and were of normal configuration with intermittent nonocclusive atheromatous plaques.

Numerous black petechial hemorrhages were appreciated on the surface of the cortex, most prominently in the left temporal lobe but also in the right temporal lobe and in the bilateral parietal and occipital lobes. Additionally, numerous punctate hemorrhages were also found on the cut surfaces of the cortex of all lobes, with the highest concentration of these being in the inferior temporal lobes. No laminar necrosis nor infarctions were found. The bilateral hippocampi were mildly atrophic.

Three blocks of tissue were submitted to Vanderbilt’s VANTAGE genomics core for DNA extraction and sequencing to assess APOE genotype. A control panel was simultaneously genotyped to ensure correct allelic discrimination. All three replicates returned identical results, homozygous C/C at rs429358 and homozygous C/C at rs7412, consistent with a homozygous E3 genotype.

### 2. Experimental methods

#### Post-mortem MR Imaging

Coronal tissue slabs were sliced coronally and embedded in 1% agarose under degassing conditions, with care to avoid air bubbles. Post-mortem susceptibility weighted imaging (SWI) and T2-weighted FLuid Attenuated Inversion Recovery (FLAIR) MRI were obtained on both 3.0T (Philips Ingenia) and 7.0T (Philips Achieva) human clinical scanners using 32-channel phased array reception. 3.0T SWI (technique=3D gradient echo, number of echoes=4, first echo=7.2 ms, echo spacing=6.2 ms, repetition time=31 ms, spatial resolution=0.6×0.6×2 mm), 3.0T FLAIR (technique=3D turbo-inversion-recovery, echo time=271 ms, repetition time=4800 ms, inversion time=1650 ms, spatial resolution=1.0×1.0×1.0 mm), 7.0T SWI (technique=3D gradient echo, number of echoes=9, first echo=8.0 ms, echo spacing=4 ms, repetition time=64.3 ms, spatial resolution=0.65×0.65×0.9 mm, and 7.0T FLAIR (technique=3D turbo-inversion-recovery, echo time=280 ms, repetition time=3952 ms, inversion time=1375 ms, spatial resolution=0.80×0.80×0.80 mm) were performed on the same day with sequence parameters chosen to parallel *in vivo* protocols.

#### Immunohistochemistry

Immunostaining of brain sections for the autopsy was conducted according to Clinical Laboratory Improvement Amendments (CLIA) standards using approved antibodies. For exploratory analysis, floating sections were prepared on a Leica 1200S vibratome with a thickness of 50 μm or 100 μm depending on the application. Meningeal tissue was gently removed from the surface of the brain, stained and mounted as a whole-mount preparation. Aggregated tau and β-amyloid were detected using thiazine red (1 μM, Chemsaves) or methoxy-XO4 (1 μM, Tocris, Minneapolis, MN), isolectin was used as a vascular marker. The primary antibodies we used were anti-PLD3 rabbit polyclonal antibody (3 μg/mL, HPA012800; Sigma-Aldrich, St. Louis, MO), anti-ATP6V_0_A_1_rabbit polyclonal antibody (10 μg/mL, NBP1-89342, Novus Biologicals), anti-CD11b chicken polyclonal antibody (10 μg/mL, MAC, AvesLabs) and anti-IBA1 mouse monoclonal antibody (5 μg/mL, 66827-1-Ig; Proteintech, Rosemont, IL). Confocal images were acquired through the Vanderbilt Cell Imaging Shared Resource (CISR) using a Zeiss LSM 710 confocal laser-scanning microscope (Carl Zeiss AG, Germany) with a 20× air/dry or 63× oil-immersion objective with a minimum resolution of 2000 × 2000 pixels.

#### CLARITY and lightsheet microscopy

For three-dimensional, lightsheet microscopy tissue blocks were incubated for 3 days in 4% paraformaldehyde (PFA) and embedded in CLARITY acrylamide hydrogel (4% Acrylamide, 0,05% Bis-Acrylamide, 0.25% temperature-triggering initiator VA-044 in 0.1 M phosphate-buffered saline in water-PBS) for X days. The tissue was then placed in a 37°C water bath to initiate hydrogel polymerization. The blocks were passively clarified in 200 mM boric acid, 4 % w/v SDS, pH 8.5 at 37°C until translucent, then pigmentation was photo-cleared with exposure to an LED microarray. The blocks were stained with a primary antibody against collagen IV (mouse anti-human, Alexa Fluor 488, concentration 1:200, Cat# 53987182 from Invitrogen) and thiazine red (Chemsaves #2150336) in PBS-Triton 0.1%. After overnight incubation in 68% thiodiethanol (TDE) to achieve a refractive index of 1.33, the blocks were imaged using a Light Sheet microscope. Image processing was done with Imaris Microscopy Image Analysis Software (Oxford Instruments).

### 3. Neuroimaging supplement

**Supplementary Figure 1:**
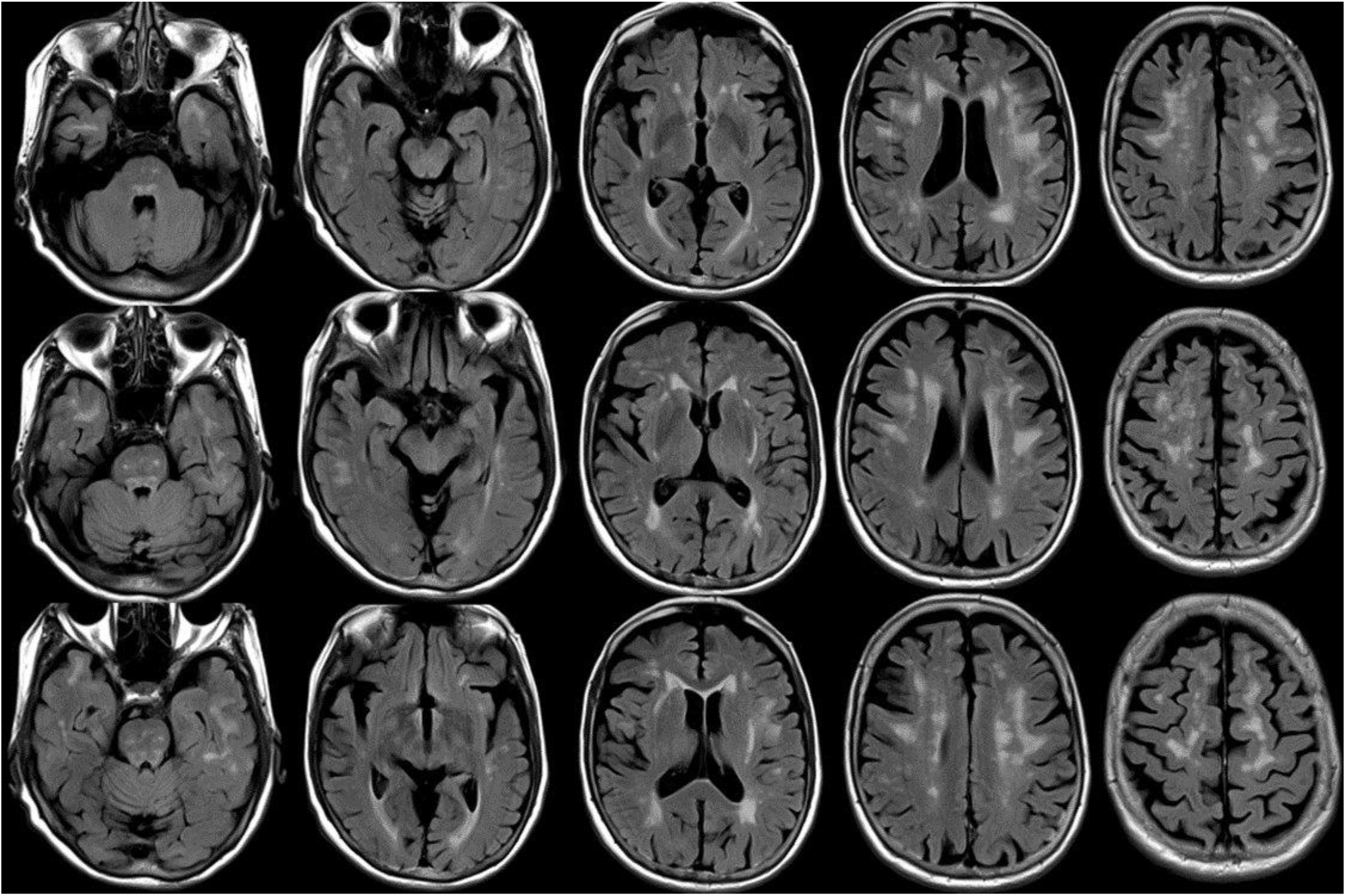
Montage of FLAIR images from pre-event MRI

**Supplementary Figure 2.**
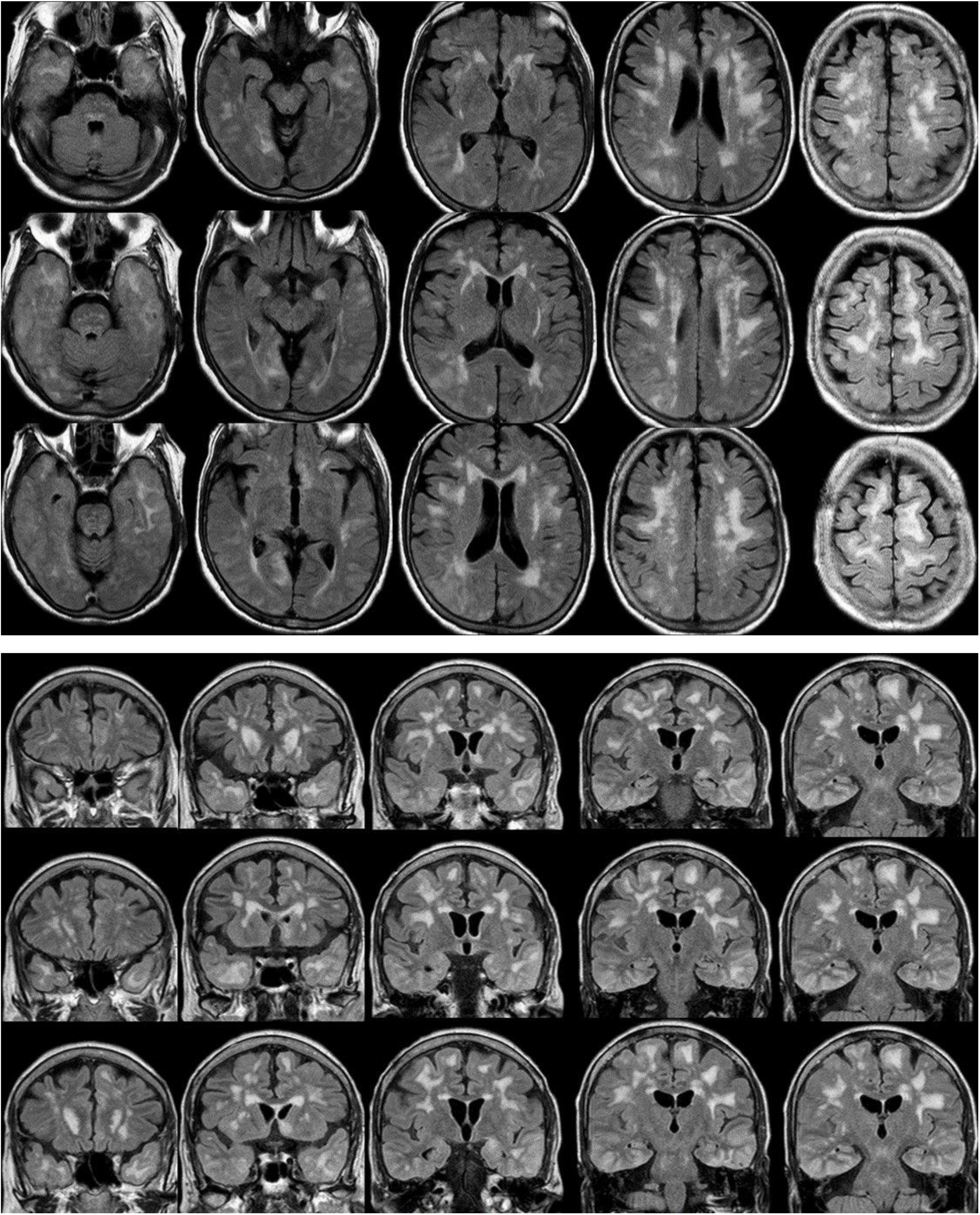
Montage of FLAIR images from post-event MRI

**Supplementary Figure 3.**
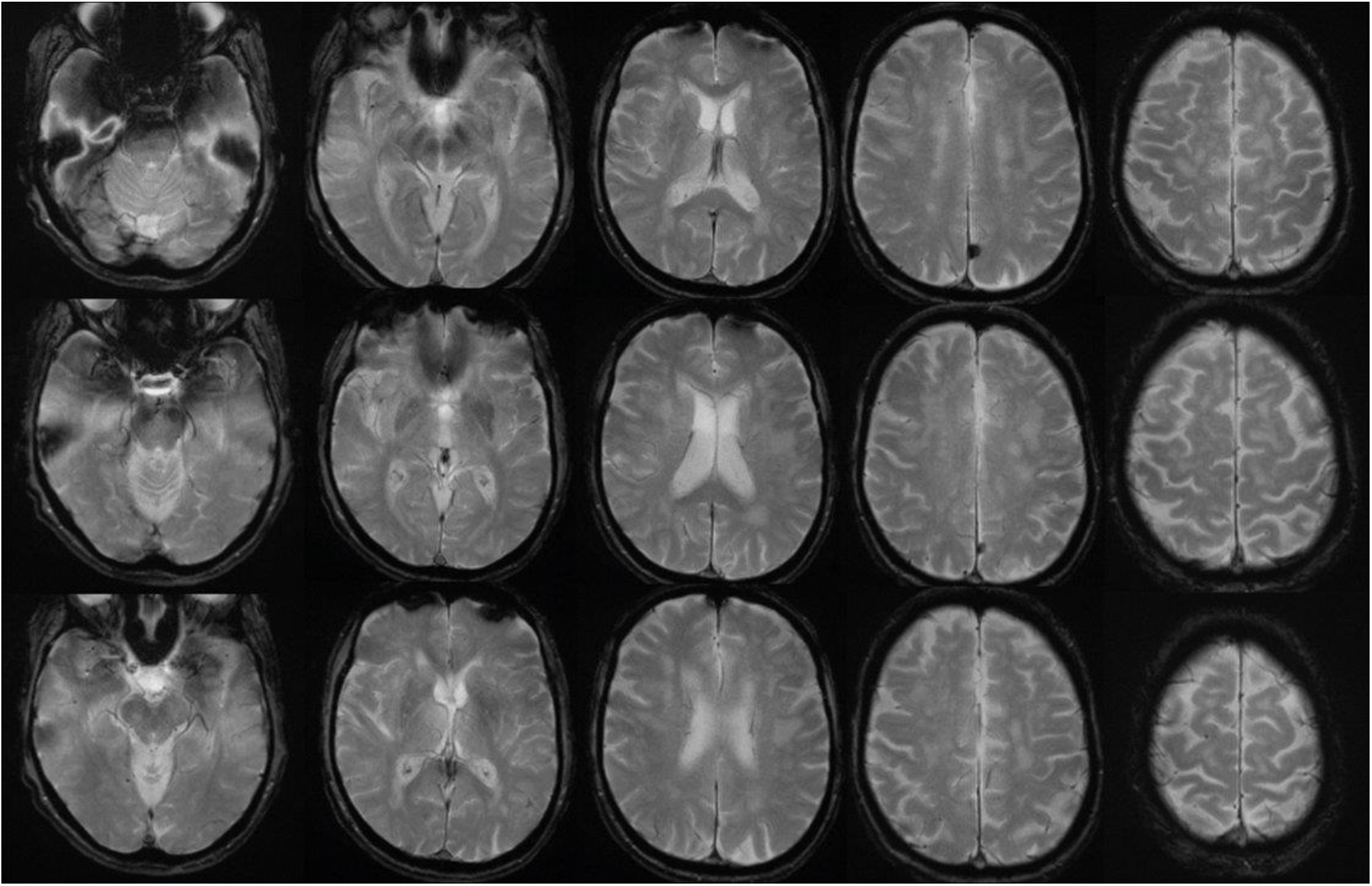
Montage of GRE-T2* images from pre-event MRI

**Supplementary Figure 4.**
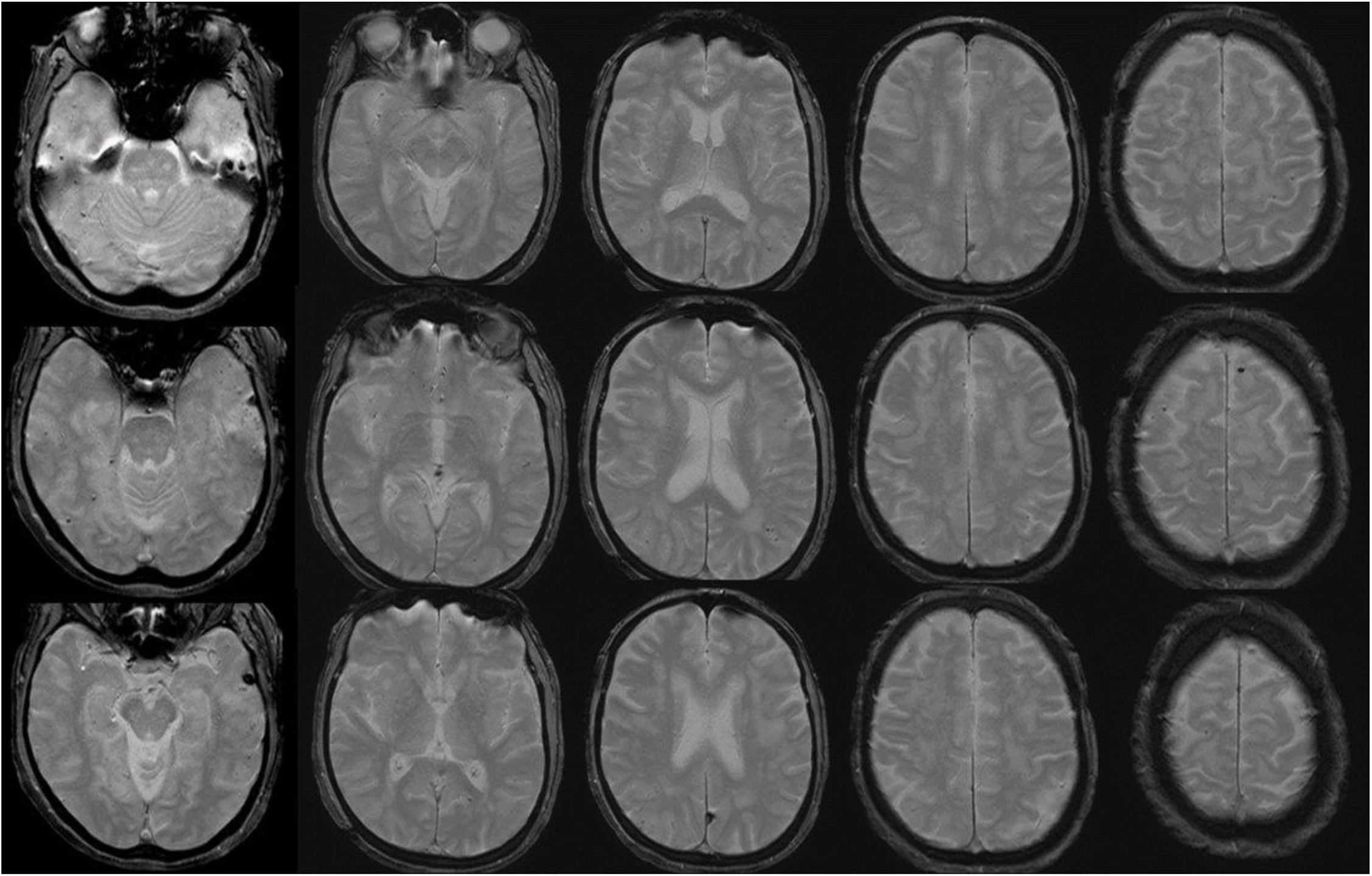
Montage of GRE-T2* images from post-event MRI

**Supplementary Figure 5.**
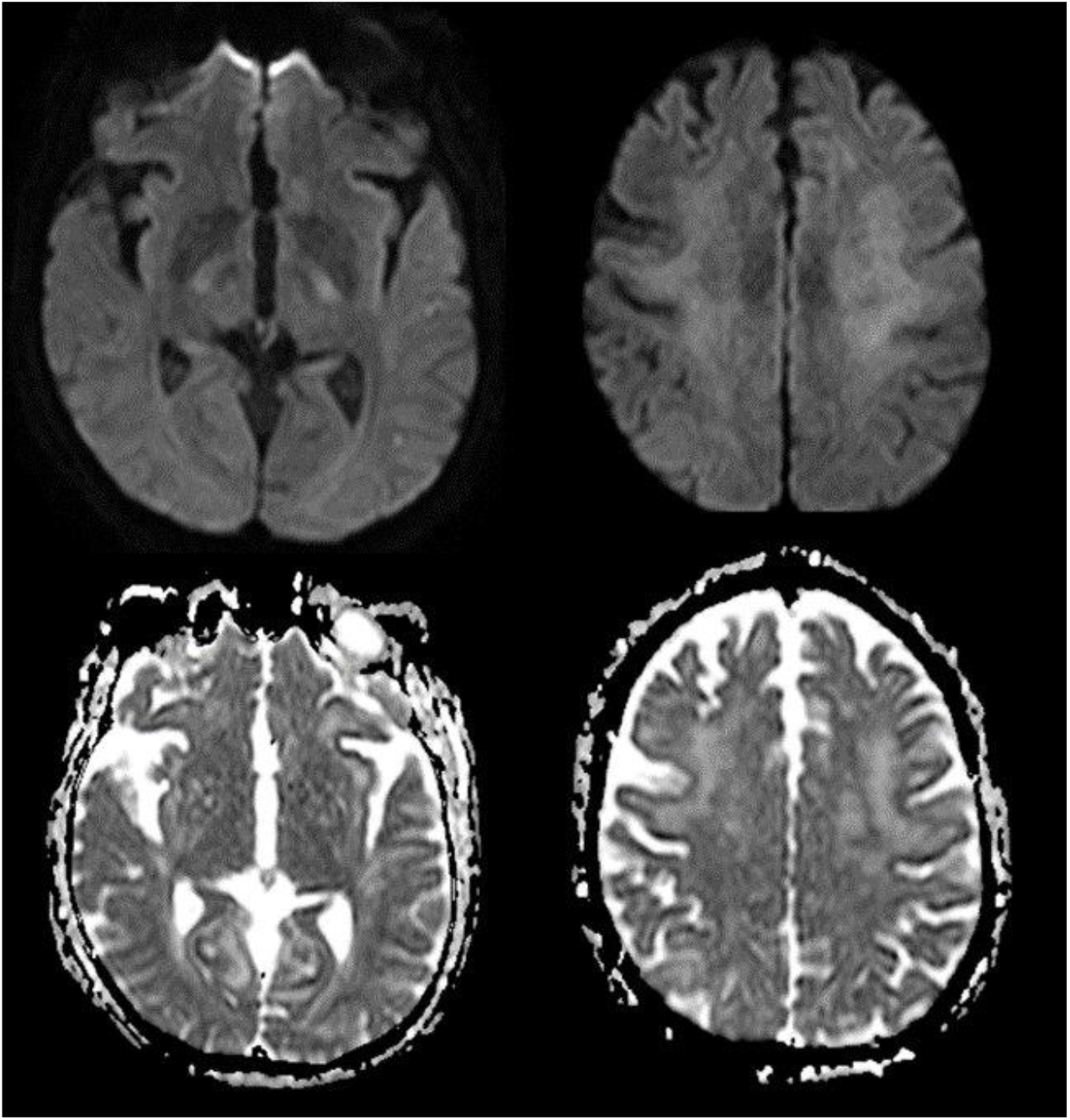
Selected images from diffusion weighted scan (hospital acquired / post-event) A. DWI and B. ADC images show no large areas of stroke. Two tiny foci of equivocal diffusion restriction are present in the left parietotemporal regions. There is also subtle cortical ribboning in insular cortices and frontal lobes.

**Supplementary Figure 6:**
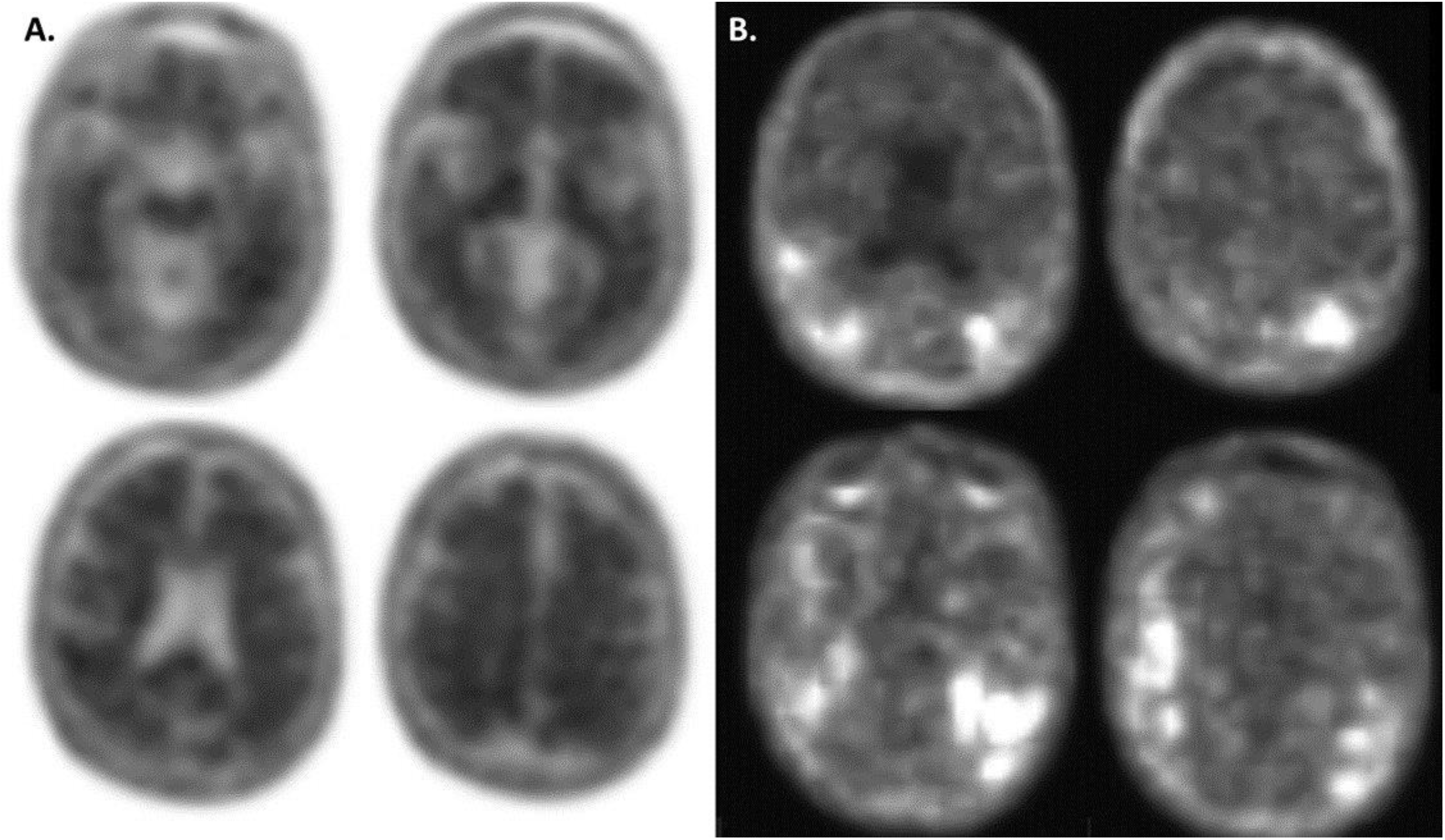
Selected images from Fluorbetaben PET and Tau PET imaging. A. Montage of selected images from Fluorbetaben PET scan obtained at enrollment in the open label extension study. B. Montage of selected images from the tau PET scan obtained at enrollment in the open label extension study.

### 4. Additional neuropathological images

**Supplementary Figure 7.**
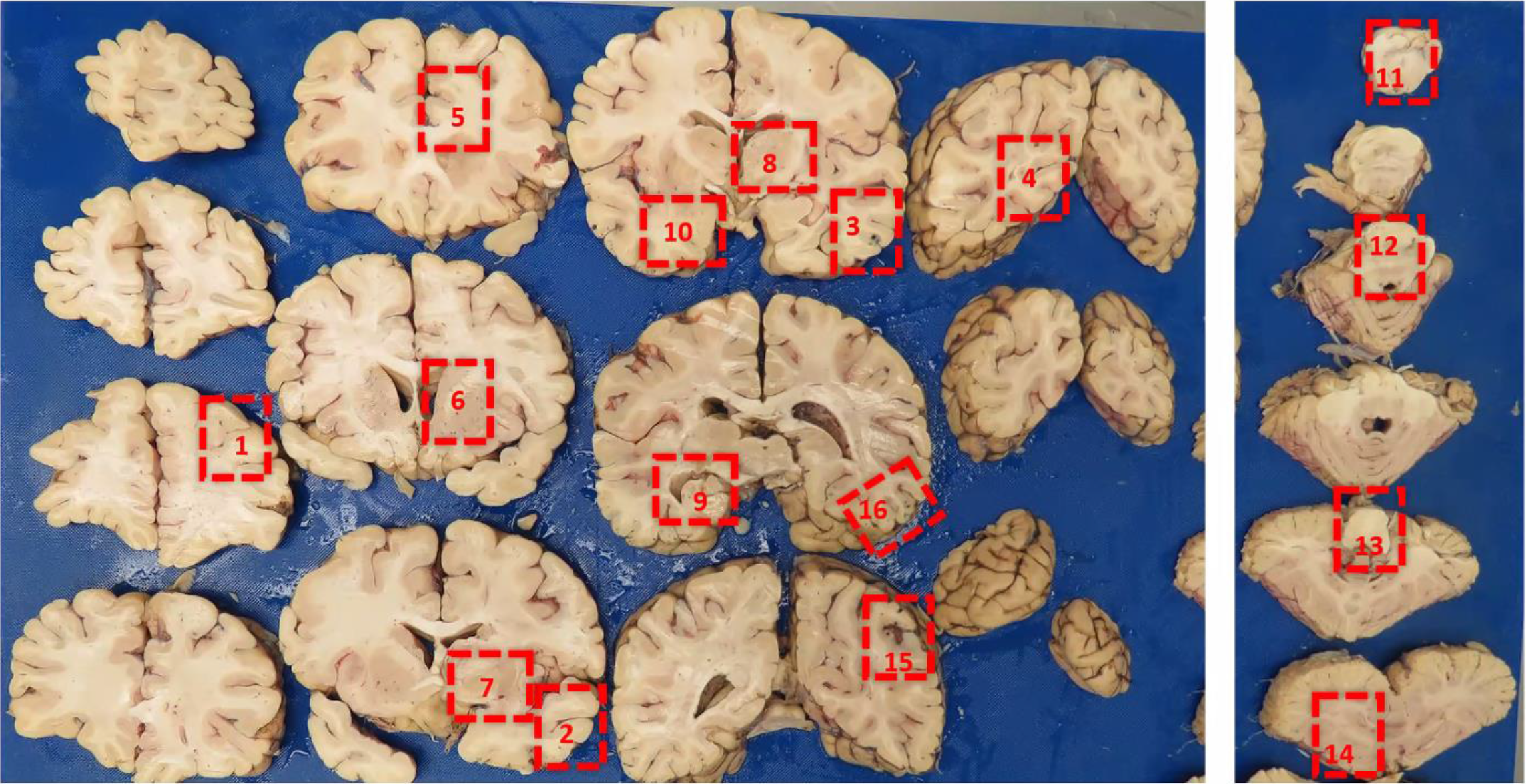
Overview image of the coronal sectioning of the brain. Red rectangles mark the tissue selected for the autopsy.

**Supplementary Figure 8:**
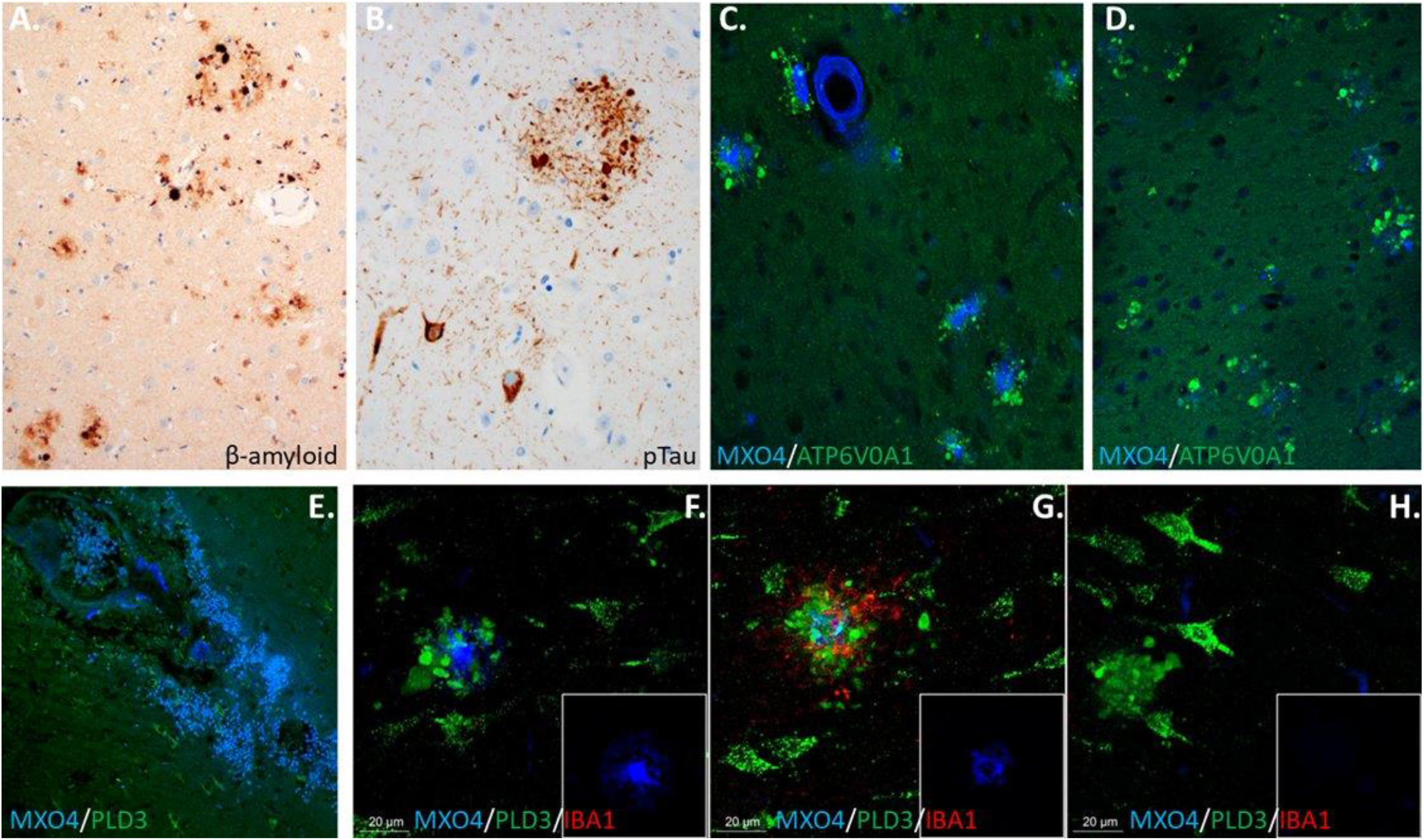
β-amyloid and tau pathology The autopsy revealed **A**. moderate density of β-amyloid plaques and **B**. a range of tau pathologies, consistent with moderate Alzheimer’s disease neuropathologic changes. **C**. This frontal lobe section shows numerous plaques and vascular amyloid deposition (stained blue with MXO4) and dystrophic neurites around plaques (stained green with the marker ATP6VOA1). **D**. This section from the medial temporal lobe shows numerous rosettes of dystrophic neurites (green), some of which lack central amyloid staining entirely and others with faint staining. **E**. A ruptured microaneurysm on a vessel with heavy amyloid deposition. **F**. A normal appearing plaque with a dense core stained brightly in blue is surrounded by a halo of dystrophic stained with the neuronal lysosome marker PLD3. **G**. A plaque that appears partially cleared (residual dense core remains present) is also surrounded by dystrophic neurites along with extensive microgliosis (microglia stained with IBA1 in red). **H**. A halo of dystrophic neurites without detectable β-amyloid is likely a site of β-amyloid clearance. Images **F-H** are z-stacks covering 15 μM in the z-plane with images every 3 μM to ensure the core of the halo of dystrophic neurites is well-captured (G is the same plaque as in figure 2d). Micrographs were obtained of 110 clusters of dystrophic neurites in the temporal lobe, hippocampus and parietal lobe, of which, 21% lacked central amyloid staining and 24% had faint residual staining.

**Supplementary Figure 9:**
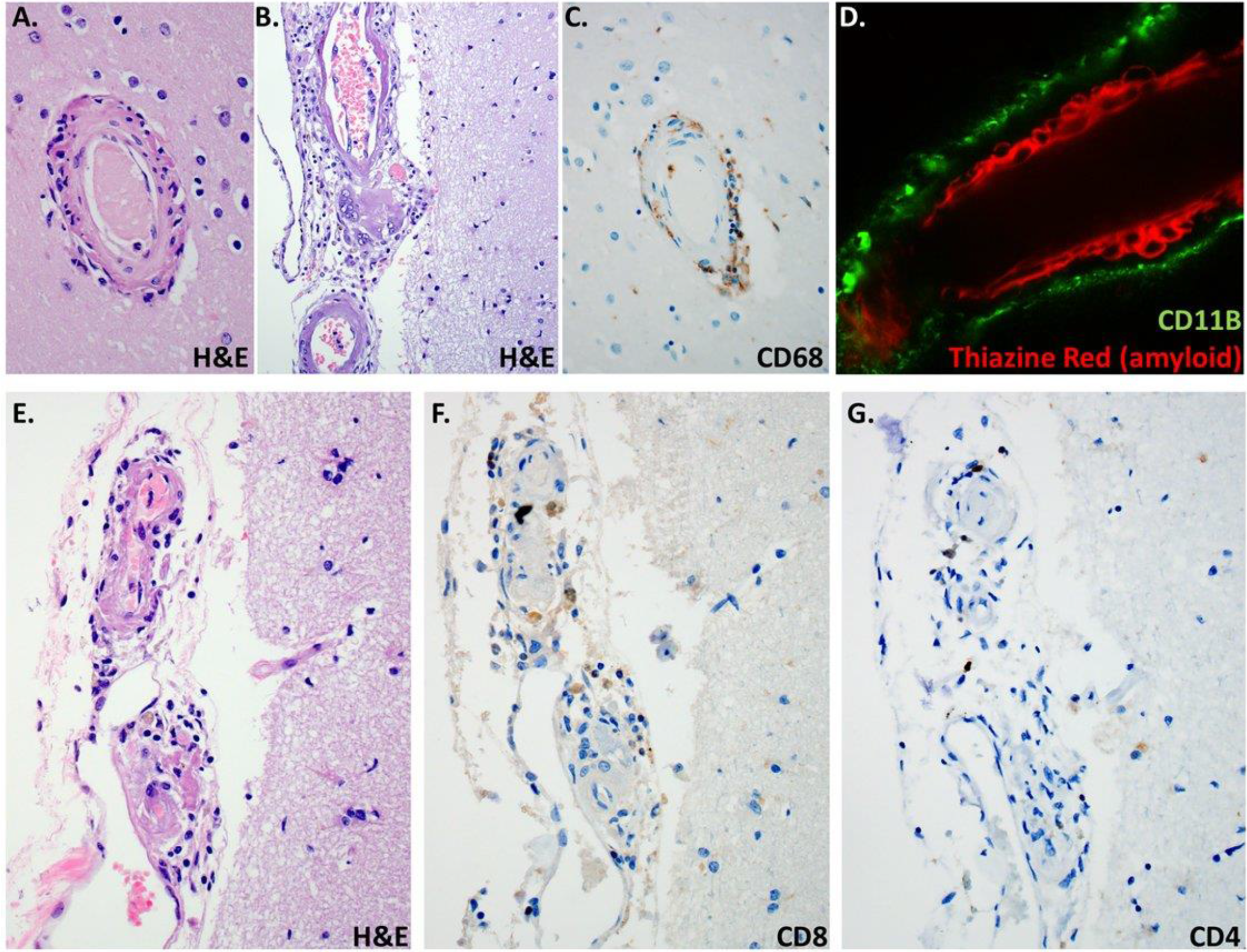
Characterization of cell types in perivascular inflammation **A**. Hematoxylin and eosin staining demonstrates inflammatory perivascular infiltrates. **B**. Multinucleated giant cells are visible in this image. **C**. CD68 immunostaining shows that many of the inflammatory cells are macrophages and/or activated microglia. **D**. CD11b, a marker of microglia and macrophages, shows numerous macrophages in the perivascular space around a vessel with heavy amyloid deposition (shown by red staining with Thiazine red). Images **E-G** show that the perivascular inflammatory infiltrate also contains T-cells, although they are less abundant than macrophages. Image **E**. shows hematoxylin and eosin staining of an inflamed vessel, **F**. CD8 staining and **G**. CD4 staining of the same vessel.

### 5. Supplementary video

*Legend for supplementary video: These three-dimensional images of arterioles associated with hemorrhages were obtained with light sheet microscopy of cleared human brain specimens. The first vessel shown is stained with thiazine red (in red, staining amyloid) and collagen 4 (in green, staining the vessel). The area of hemorrhage appears somewhat yellow due to autofluorescence. The second vessel is stained similarly. The pink channel shown at the end of the video shows only autofluorescence (no primary antibody), highlighting the location of blood. In both ruptured vessels deposits of amyloid are present at the site of rupture*.

## Notes

### Funding Statement

This study was made possible by philanthropic support from the family and friends of Louis Stephen Zuzga Moran and the family and friends of Douglas B. Janney, Jr.

### Author Declarations

Vanderbilt University Medical Center's institutional review board granted ethical approval of this study as part of our ongoing Observational Study of CAA and Related Disorders (OSCAAR), approval number 180287.

## References

1. Birmingham K, Frantz S. Set back to Alzheimer vaccine studies. Nat Med. 2002;8(3):199–200.

2. Nicoll JAR, Buckland GR, Harrison CH, Page A, Harris S, Love S, et al. Persistent neuropathological effects 14 years following amyloid-beta immunization in Alzheimer’s disease. Brain. 2019;142(7):2113–26.

3. Auriel E, Charidimou A, Gurol ME, Ni J, Van Etten ES, Martinez-Ramirez S, et al. Validation of Clinicoradiological Criteria for the Diagnosis of Cerebral Amyloid Angiopathy-Related Inflammation. JAMA Neurol. 2016;73(2):197–202.

4. Budd Haeberlein S, Aisen PS, Barkhof F, Chalkias S, Chen T, Cohen S, et al. Two Randomized Phase 3 Studies of Aducanumab in Early Alzheimer’s Disease. J Prev Alzheimers Dis. 2022;9(2):197–210.

5. Nackenoff AG, Hohman TJ, Neuner SM, Akers CS, Weitzel NC, Shostak A, et al. PLD3 is a neuronal lysosomal phospholipase D associated with beta-amyloid plaques and cognitive function in Alzheimer’s disease. PLoS Genet. 2021;17(4):e1009406.

6. Schrag M, McAuley G, Pomakian J, Jiffry A, Tung S, Mueller C, et al. Correlation of hypointensities in susceptibility-weighted images to tissue histology in dementia patients with cerebral amyloid angiopathy: a postmortem MRI study. Acta Neuropathol. 2010;119(3):291–302.

7. Sperling RA, Jack CR, Jr., Black SE, Frosch MP, Greenberg SM, Hyman BT, et al. Amyloid-related imaging abnormalities in amyloid-modifying therapeutic trials: recommendations from the Alzheimer’s Association Research Roundtable Workgroup. Alzheimers Dement. 2011;7(4):367–85.

8. Scheltens P, Goos JD. Dementia in 2011: Microbleeds in dementia--singing a different ARIA. Nat Rev Neurol. 2012;8(2):68–70.

9. Schindler RJ, Carrillo MC. Output of the working group on magnetic resonance imaging abnormalities and treatment with amyloid-modifying agents. Alzheimers Dement. 2011;7(4):365–6.

10. Salloway S, Chalkias S, Barkhof F, Burkett P, Barakos J, Purcell D, et al. Amyloid-Related Imaging Abnormalities in 2 Phase 3 Studies Evaluating Aducanumab in Patients With Early Alzheimer Disease. JAMA Neurol. 2022;79(1):13–21.

11. Nicoll JA, Wilkinson D, Holmes C, Steart P, Markham H, Weller RO. Neuropathology of human Alzheimer disease after immunization with amyloid-beta peptide: a case report. Nat Med. 2003;9(4):448–52.

12. Ferrer I, Boada Rovira M, Sanchez Guerra ML, Rey MJ, Costa-Jussa F. Neuropathology and pathogenesis of encephalitis following amyloid-beta immunization in Alzheimer’s disease. Brain Pathol. 2004;14(1):11–20.

13. Liu E, Wang D, Sperling R, Salloway S, Fox NC, Blennow K, et al. Biomarker pattern of ARIA-E participants in phase 3 randomized clinical trials with bapineuzumab. Neurology. 2018;90(10):e877–e86.

14. Charidimou A, Boulouis G, Frosch MP, Baron JC, Pasi M, Albucher JF, et al. The Boston criteria version 2.0 for cerebral amyloid angiopathy: a multicentre, retrospective, MRI-neuropathology diagnostic accuracy study. Lancet Neurol. 2022;21(8):714–25.

15. Knudsen KA, Rosand J, Karluk D, Greenberg SM. Clinical diagnosis of cerebral amyloid angiopathy: validation of the Boston criteria. Neurology. 2001;56(4):537–9.

16. Sevigny J, Chiao P, Bussiere T, Weinreb PH, Williams L, Maier M, et al. The antibody aducanumab reduces Abeta plaques in Alzheimer’s disease. Nature. 2016;537(7618):50–6.

17. Heuer E, Jacobs J, Du R, Wang S, Keifer OP, Cintron AF, et al. Amyloid-Related Imaging Abnormalities in an Aged Squirrel Monkey with Cerebral Amyloid Angiopathy. J Alzheimers Dis. 2017;57(2):519–30.

18. Zago W, Schroeter S, Guido T, Khan K, Seubert P, Yednock T, et al. Vascular alterations in PDAPP mice after antiAbeta immunotherapy: Implications for amyloid-related imaging abnormalities. Alzheimers Dement. 2013;9(5 Suppl):S105–15.

19. Alves F, Kallinowski P,Ayton S. Accelerated Brain Volume Loss Caused by Anti-beta-Amyloid Drugs: A Systematic Review and Meta-analysis. Neurology. 2023.

